# Epidemiology of Relapsing and Falciparum Malaria in the Highlands of Cameroon: An Integrated Community Survey of Human Infection and Vector Abundance

**DOI:** 10.1101/2025.04.28.25326551

**Authors:** Samuel J. White, Valery P. K. Tchuenkam, Mariama Mbouh, Claudia Gaither, Aline Gaelle Bouopda-Tuedom, Belinda Kiam, Zachary R. Popkin-Hall, Jacob M. Sadler, Kelly Carey-Ewend, Emily Hand, Miriam N. Ngum, Yannick N. Ngomsi, Jeffrey A Bailey, Darlin B. Kaunda, Lethicia K. Mafo, Giresse N. Lemogo, Clifford L. Dinka, Clifford A. Nsani, Michel Noubom, Varun Goel, Ibrahima Ibrahima, Janvier C. Onguene, Feng-Chang Lin, Jessica T. Lin, Sandrine E. Nsango, Rhoel R. Dinglasan, Jonathan J. Juliano, Innocent M. Ali

## Abstract

Despite global malaria control efforts, the disease caused 263 million cases and 597,000 deaths in 2023. While *Plasmodium falciparum* accounts for most cases in Africa, non-falciparum species, such as *P. ovale* spp. and *P. vivax*, can cause relapse infections and are increasingly recognized as significant contributors to human disease. In particular, the highlands of West Cameroon have previously been reported to have high *P. vivax* infection rates. This study presents preliminary results from the Relapsing Malaria in Africa (ReMA) study, conducted in Dschang, Cameroon, to assess the prevalence and epidemiology of *P. vivax* and *P. ovale*. A cross-sectional survey of 3,661 participants from 871 households across seven health areas identified a low prevalence of *P. vivax* (0.1%) and *P. ovale* spp. (0.64%) using quantitative real time PCR (qPCR), while *P. falciparum* remained prevalent at 8.1%. Co-infections of *P. ovale* spp. with *P. falciparum* were observed in 23.1% of *P. ovale* spp. cases. While gametocytemia was common among falciparum infections, leveraging a new ovale gametocyte assay, we found that gametocytemia was uncommon among the qPCR-positive ovale infections. Spatial analysis found *P. vivax* and *P. ovale* spp. infections concentrated in Penka-Michel and Baleveng, the former having higher *Anopheles* spp. abundance than other areas assessed. Risk factor analysis revealed children and those with recent domestic travel had higher odds of *P. falciparum* infection, but no significant associations were found for *P. ovale* spp. infections. Entomological surveys confirmed high densities of *Anopheles gambiae sensu lato* (*s.l.*) and *An. funestus* (s.l.), with significantly higher human landing capture rates for *An. gambiae* s.l compared to other mosquito species. While these findings suggest that the relapsing malarias are not as widespread as previously thought in West Cameroon, understanding factors driving their persistent transmission, especially without prevalent gametocytemia, will be important for disease control.

**AUTHOR SUMMARY:** Malaria caused 263 million cases and 597,000 deaths in 2023, with most cases in Africa due to *Plasmodium falciparum*. However, the dangers of other species like *P. ovale* and *P. vivax*, which can cause relapsing infections, are becoming more clear in Africa. The Relapsing Malaria in Africa (ReMA) study in Dschang, Cameroon, surveyed 3,661 people from 871 households to better understand these infections. Results showed low rates of *P. vivax* (0.1%) and *P. ovale* (0.64%) compared to *P. falciparum* (8.1%). Infections were more common in areas with high mosquito abundance and biting activity. While the relapsing malarias were less common than expected, understanding factors driving their persistent transmission, especially without prevalent gametocytemia, will be important for disease control.

## INTRODUCTION

Despite global control efforts, there were 263 million malaria cases and 597,000 deaths from malaria in 2023 (1). While most of the burden is caused by *Plasmodium falciparum* in Africa, there is growing evidence that non-falciparum malaria is becoming more common (2–4). Three species, *P. ovale* spp., consisting of *P. ovale curtisi* and *P. ovale wallikeri*, as well as *P. vivax* cause relapsing malaria, which further complicate control by requiring radical cure for dormant hypnozoites. The assumption that *P. vivax* and *P. ovale* spp. results in only benign infections have also been challenged by increased reports of severe disease, particularly in young children and pregnant women (5–8). *P. ovale* infections are known to be endemic to Africa, but an increase in prevalence has been observed in some countries in the face of declining *P. falciparum* infections (4,9–11). Historically, *P. vivax* has been assumed to be largely absent from Sub-Saharan Africa due to the predominance of the Duffy-negative phenotype in Sub-Saharan African populations (12). However, cases of *P. vivax* have been repeatedly documented in studies showing *P. vivax* infection in Duffy-negative populations across Africa (13–17).

*P. ovale* spp. and *P. vivax* pose unique challenges to elimination efforts in Africa. Due to distinct biological characteristics, including early development of gametocytes and liver dormancy as hypnozoite stages, risk factors for these species typically do not match those for *P. falciparum* in Africa (18). They also tend to occur in lower transmission settings in Africa (9,19–21). The undetectable hypnozoite stage and the typically low asexual parasitemia of *P. ovale* spp. and *P. vivax* infections in Africa relative to *P. falciparum* make diagnosis by microscopy difficult even among trained microscopists (22). Molecular methods such as polymerase chain reaction (PCR) are more sensitive, particularly for asymptomatic infections, and allow for the detection of mixed infections.

Relapsing malarias have been detected in Cameroon, but with inconsistent findings surrounding their epidemiology (**Table 1**). Almost all previous studies of the relapsing malaria in Cameroon have been in febrile clinical populations. Outside of the West province, overall prevalence of *P. ovale* spp. and *P. vivax* has been low, never encompassing more than 5% of screened individuals (23–29). These studies were done in geographically diverse regions of Cameroon, and represented each of the five distinct ecological zones of Cameroon between them (Sahelian, Soudanian, Sahelo-Guinean, Humid Savannah and Forest) (30). Recent studies in the West region, around Dschang, have identified these species more commonly in febrile clinical patients. Russo et al. found *P. vivax* infection in 5.6% of febrile outpatients in 2017, while Dongho et al. reported that 35.2% of febrile patients in the hospital were PCR positive for *P. vivax* during the rainy season in 2021 (14,31). Previous work by our group in 2020 found 7% of malaria positive individuals who presented to clinics had *P. ovale* spp. infection (29). Given the significant number of *P. ovale* spp. and *P. vivax* infections detected in clinics in the region, a deeper understanding of relapsing malaria epidemiology is needed in the Western highlands; yet no comprehensive community survey has been completed.

**Table 1:** Previous studies of relapsing malaria in Cameroon.

| Citation | Sampling period | Region(s) | Setting | PCR positive for *P. ovale* spp. | PCR positive for *P. vivax* |
| --- | --- | --- | --- | --- | --- |
| Fru-Cho et al. 2014 | 2008-2009 | Southwest | Clinic | 0/269 (0%) | 13/269 (4.8%) |
| Mbenda and Das 2014 | 2012 | Littoral, Centre, South and East | Clinic | 0/485 (0%) | 8/485 (1.6%) |
| Russo et al. 2017 | 2016 | West | Clinic | 0/484 (0%) | 27/484 (5.4%) |
| Kwente et al. 2017 | 2017 | Far North, Centre, Northwest, Littoral and Adamawa | Clinic | 0/1609 (0%) | 0/1609 (0%) |
| Dongho et al. 2021 | 2017 | West and South | Clinic | 2/1001 (0.2%) | 142/1001 (14.2%) |
| Foko et al. 2022 | 2019 | Far North, North and Littoral | Clinic | 3/98 (3%) | 1/98 (1%) |
| Sofeu-Feugaing et al. 2023 | 2020 | Littoral, Southwest, West and Centre | Clinic | 26/798 (3.3%) | 0/798 (0%) |
| Apinjoh et al. 2024 | 2019-2021 | Far North, Adamawa, Northwest, Southwest, Littoral, Center and East | Clinic and community | 73/1785 (4.1%) | 0/1786 (0%) |
| Tchuenkam et al. 2025 | 2020 | West | Clinic | 17/431 (3.9%) | 0/431 (0%) |

*Plasmodium* spp. are transmitted by anopheline vectors, which are highly diverse in Cameroon with at least 60 known species (32). Currently, long-lasting insecticidal nets (LLIN) are the only vector control strategy used by the National Malaria Control Program of Cameroon, but insecticide resistance has been observed in *An. gambiae sensu lato* (*s.l.*) and *An. funestus* (*s.l.*), primarily to dichlorodiphenyltrichloroethane (DDT), permethrin, deltamethrin and bendiocarb (33). Previous entomological studies in the West Region of Cameroon have found *An. gambiae* (*s.l.*) and *An. funestus* (*s.l.*) to be the primary *P. falciparum* malaria vectors, but *An. nili* (*s.l.*), *An. ziemanni,* and *An. rufipes* have also been reported (34,35). Despite these efforts, the primary vectors for *P. ovale* and *P. vivax,* which included *An. coluzzii* previously in Cameroon remain unclear (36).

To provide integrated community-based surveys of malaria and vectors in the Western highlands, the Relapsing Malaria in Africa (ReMA) study was launched in 2023 to better describe the epidemiology and transmission of relapsing malaria and falciparum malaria in Dschang and the surrounding area in the West Region, Cameroon. This report details the results of the preliminary cross-sectional community survey, which aimed to: 1) estimate *P. ovale* spp. and *P. vivax* prevalence in the community, 2) determine the rate of *P. ovale and P. vivax* gametocytemia, 3) determine geographical distribution of different *Plasmodium* species, 4) define risk factors associated with *P. ovale* spp. and *P. vivax* infection in the region, and 5) determine vector species and abundance in the communities relative to household collection of asymptomatic infections.

## METHODS

### Ethics Statement

Signed written consent was obtained by all adult participants (>20 years of age) enrolled in the study. Written, signed parental consent and assent from participants 20 years of age or younger was also provided. Children under the age of 6 years old were not eligible for enrollment, as components of the overall project not described here involved the need to give meaningful assent, which the IRBs deemed not possible for those under 6 years of age. The study protocol was approved by the Cameroon National Ethics Committee for Human Health Research (N°2022/12/1507/CE/CNERSH/SP) and University of North Carolina at Chapel Hill (22–1445).

### Study site

A cross-sectional community survey was performed in the city of Dschang, in the Fialah Foréké, Siteu, Fometa, and Doumbouo neighborhoods, and surrounding areas [Baleveng, Mboua and Penka-Michel (Centre Urbain)], located in the West Region of Cameroon (**Figure 1**). These sites were identified as having higher malaria transmission using data from the 2022 Dschang Health District annual malaria report. Specific characteristics about malaria and environmental characteristics that impact malaria transmission for each health area are shown in **Table 2**, including rapid test positivity rate from health center data, estimates of vegetation coverage, and percentage of built space (as a proxy of urban development). Malaria positivity by RDT in health center data ranged from 39.7% in Siteu to 72.2% in Doumbouo, with built-up land percentage ranging from 3.0% in Mboua and 23.9% in Fialah Foréké.

**Figure 1.**
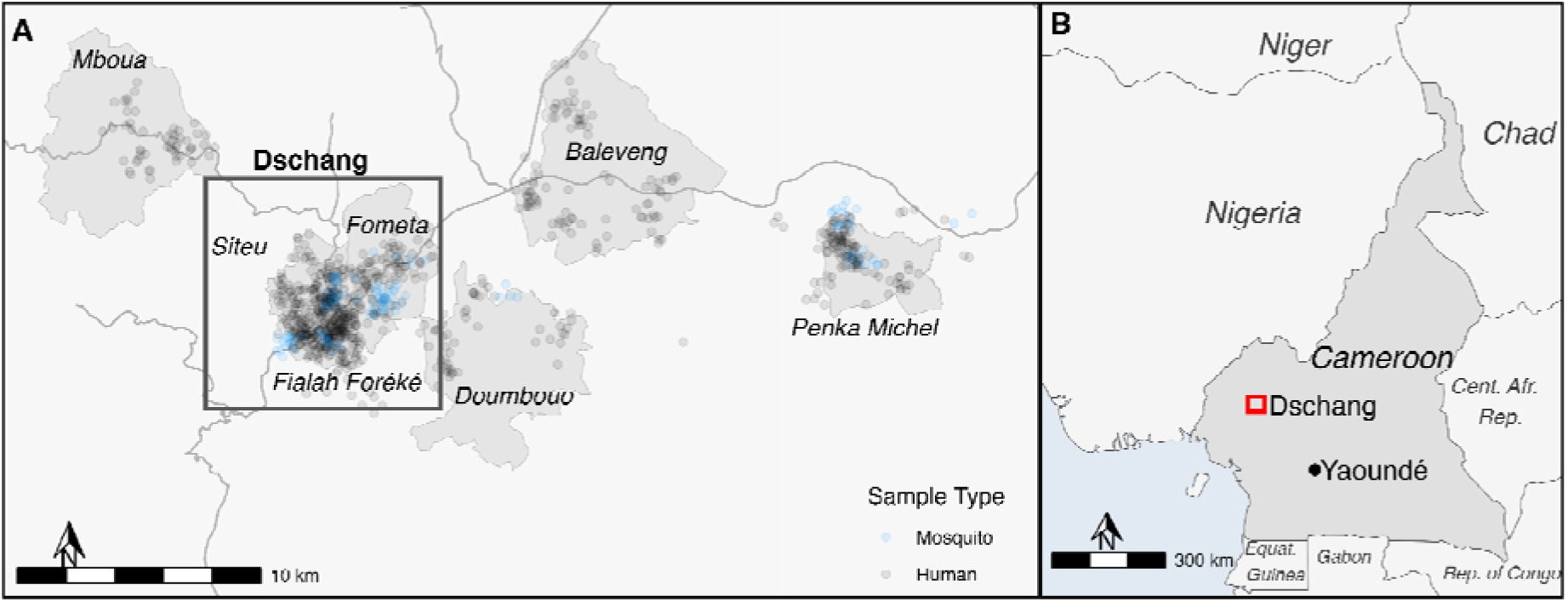
Map of study area and distribution of households sampled and mosquito collections. In Panel A, light grey areas represent administrative boundaries. Household and mosquito sampling locations are each randoml jittered a distance of 450m. The region sampled is highlighted by the red box in Panel B.

**Table 2.** Study Sites Characteristics (May-Sep 2023)

| Site name | Population(37) | NMCP reported malaria cases/tested in July 2023<sup>#</sup> | Vegetation Index<sup>%</sup> | Built-up Land(<sup>%</sup>)<sup>^</sup> |
| --- | --- | --- | --- | --- |
| Fialah Foréké | 42,925 | 61.6% | 0.54 | 23.9% |
| Fometa | 19,524 | 71.6% | 0.69 | 12.3% |
| Siteu | 28,653 | 39.7% | 0.63 | 17.7% |
| Doumbouo | 12,950 | 72.2% | 0.75 | 5.7% |
| Penka Michel-Centre Urbain | 19,890 | 68.5% | 0.73 | 7.6% |
| Mboua | 8,003 | 52.5% | 0.79 | 3.0% |
| Baleveng | 19,198 | 50.0% | 0.74 | 7.1% |

<sup>#</sup>: Data on malaria from NMCP, total number of tests per site is unavailable..
<sup>%</sup>: Data from MODIS Terra calculated as the mean NDVI between May 1- Sep 30, 2023 for the study site; Higher values suggest denser vegetation.
<sup>^</sup>: Data from Global Human Settlement Layer 2020. Calculated as percentage of total built-up area within the study site boundary

Dschang, the capital of the Menoua division of the West Region, has a population of roughly 130,000. It is in a humid savannah approximately 1380 meters above sea level and receives 2000mm annual rainfall. Temperatures range from a low of 13.6°C to a high of 25.3°C throughout the year (mean 20-23°C) and the region has two distinct seasons: 1) the main rainy season (mid-March to early October), which is punctuated by a short dry season in July, and 2) the main dry season (November to early March). The area is considered hypoendemic for malaria and has seasonal malaria transmission lower than much of the rest of the country (30). The primary industry in Dschang is agriculture but there is a large student population (14,000 full-time students) that attend the University of Dschang. The peri-urban neighborhoods of Dschang are mostly made up of permanent housing and are dominated by hills with an average elevation of 1450m above sea level. Dschang also includes multiple small lakes in the urban centre and marshy lowlands with creeks and areas around watercourses used for market gardening and backyard pig rearing. Most of the sampled neighborhoods/villages outside of Dschang consist of an urban center surrounded by rural areas with a primarily agricultural population. The landscape is further characterized by a succession of high peaks and deep valleys (Doumbouo) or an alternate series of undulating landforms. Baleveng shares similar features and lies along the regional road linking Bafoussam, the regional capital, and Dschang. Penka-Michel on the other hand has a hilly relief punctuated with savannas and forests, and small streams that traverse the neighbourhood of Centre Urbain, a semi-rural health area in the Penka-Michel health district. The altitude of these neighbourhoods vary between 1300m-1450m.

### Household sampling and enrollment

Sampling occurred during the latter part of the main rainy season from May 10th to June 19th, and August 4th to September 18th, 2023. Within the selected health areas, a random selection of clusters, representing neighborhoods in urban areas and villages in rural areas, were identified. Sixty households were targeted to be sampled per cluster. When possible, the first household visited in each neighborhood was the local chief’s and then a bottle was spun to pick a direction to continue sampling of further households in that direction. Sampling weights were calculated for each cluster by taking the inverse of the product of the probability of a cluster being selected and the probability of a household in that cluster being selected (**Supplemental Table 1**). If a sampled household was not available to be surveyed, the neighboring household was visited. Population and household distribution estimates used to construct sampling weights were taken from the 2022 National Onchocerciasis Control Program (PNLO) (37). The distribution of households and vector sampling sites is shown in Figure 1.

### Survey and sample collection

Each participant was interviewed with responses recorded on a tablet using REDCap (Research Electronic Data Capture) (38,39). Demographic information, education level, occupation, travel history, and malaria diagnosis and treatment history were recorded for each participant. The temperature of each participant was recorded. If febrile, they were tested for malaria with a P.f/Pan malaria rapid diagnostic test (RDT) (Abbott Laboratories, Green Oaks, Illinois). For each participant, dry and liquid blood spots were collected from a finger-prick, and hemoglobin concentration was measured (Hemocue 801, Ängelholm, Sweden). Dried blood spots (DBS) were collected on Whatman 3MM filter paper, dried, and stored in packets with desiccant (Cytiva, Marlborough, Massechussetts). Whole blood was collected into microfuge tubes containing 2X DNA/RNA shield (Zymo Research, Irvine, CA) (40). The head of each household was given an additional interview on family size, bed net use, previous malaria treatment and its monetary cost, livestock ownership, household water source, and housing material.

### Detection of human *Plasmodium sp.* infection

Parasite DNA was extracted from DBS using Extracta DBS (QuantaBio, Beverly, MA) following the manufacturer’s protocol. Parasitemia was determined by semi-quantitative real-time PCR (qPCR) for *P. falciparum*, *P. vivax* and *P. ovale*. using species-specific 18S rRNA assays run to 45 cycles (**Supplemental Table 2**) (9,15,41). For quantification, known dilutions of plasmids (MRA-177, 178, and 180; BEI Resources, Manassas, VA) were used as previously described (3). Six copies of the species specific plasmid were used to estimate a genomic equivalent for all species (42). Each PCR plate also contained two negative controls (water). All qPCR was done at University of Dschang using a Biorad CFX96 real time PCR machine.

### Determination of gametocyte carriage

RNA from samples that were qPCR positive for *P. falciparum* and *P. ovale* spp. were extracted from up to 75 microliters of whole blood stored in DNA/RNA shield (Zymo Research) using a magnetic bead based protocol (Quick-RNA MagBead, Zymo Research). Successful RNA extraction was confirmed by detecting human-beta tubulin RNA in the first few samples using a reverse-transcriptase quantitative PCR (rt-qPCR) assay (43). *P. falciparum* gametocytes were detected as previously described using primers specific to *P. falciparum* surface protein 25 (*pfs25*) and *P. falciparum* male gametocyte-enriched transcript (*pfmget)* targets for female and male gametocytes, respectively (**Supplemental Table 3**) (44,45). Control reactions containing no reverse transcriptase were run for all *pfs25* positive samples, to ensure positivity was not due to genomic DNA amplification. This was not needed for the *pfmget* assay, as primers were designed to span an intron. Positive controls for falciparum gametocyte assays were generated by PCR amplification of malaria genomic DNA using *pfs25* and *pfmget* primers followed by cloning into plasmid vectors and transformation into *E. coli* using LB agar plates and competent cells (TOPO™ TA Cloning™ Kit, with pCR™2.1-TOPO™, Invitrogen, Waltham, Massachusetts). A conversion factor of 87.5 and 12.5 for *pfs25* and *pfmget* was used to go from transcript copies to gametocyte count, respectively.

*P. ovale* spp. female gametocytes were detected using the previously described *pos25* target (**Supplemental Table 4**), orthologous to the *pfs25* target (46). No reverse transcriptase controls have been previously described for the *pos25* target, and were not run in this analysis. Positive controls for the assays were generated using plasmid containing the *pos25* 100 bp target sequence. Transcript copies are reported for *P. ovale* assays as no conversion factor is available.

A new *P. ovale* male gametocyte assay was developed to: 1) have a single assay to detect both species and 2) utilize a gene with an intron to reduce false positive detection. The target was identified by searching for *P. ovale* primer sequences targeting the ortholog to the falciparum gametocyte target *pfmget*. Primers and probes were designed to be approximately 20 bp and intron-spanning using Primer3 (**Supplemental Table 4**) (47). Cycling conditions are described in **Supplemental Table 4**. A positive control for the ovale assay was synthesized using species-specific sequences (GeneWiz, South Planefield, NJ) to produce control plasmids.

To determine the limit of detection (LOD) for the *P. ovale* spp. gametocyte assay, probit analysis was conducted on replicate rt-qPCR runs using templates with known copy number concentrations (48). Positivity was based on a cycle threshold <45 cycles. Given that the number of transcripts for each gene per gametocyte is not known, we report on transcript copies, rather than gametocytemia. All positive samples were confirmed by manually reviewing the amplification curves using machine software. Standard curves generated from plasmid positive controls were required to have a minimum r-squared value of 0.95 for each run.

### Analysis of spatial clustering of malaria infection

To investigate the spatial distribution of malaria across the study area we estimated kernel-smoothed spatial relative risk surfaces for *P. falciparum* and *P. ovale* spp. (49). Spatial relative risk is calculated as a ratio of the kernel-estimated density of PCR positive cases and PCR negative cases, using an adaptive kernel bandwidth-estimation. It describes the risk of malaria at a particular location compared to the expected risk across the study area after taking the background population in account (50,51). We also identified clusters of malaria prevalence, defined as areas with significantly elevated malaria risk, by calculating adaptive tolerance contours at a significance level of 0.05 using 999 monte-carlo simulations (52). All spatial clustering analysis was conducted using the ‘sparr’ package in R version 4.2.2. (53).

### Mosquito abundance data

To evaluate the relationship between vector abundance and malaria cases, mosquito abundance data was determined from collections performed in these areas. Details on mosquito capture and entomological indicators are described previously (54). Here, we provide a subset analysis of these data for only those areas where community collections from participants occurred. We included these data as the abundance of mosquitoes and the rate of infection are closely linked, as larger, competent mosquito vector populations may contribute to a rise in parasite transmission.

### Statistical analysis

For categorical variables, frequencies and percentages were calculated. Continuous variables were summarized with median (IQR) unless otherwise specified. Weighted prevalences and 95% Confidence Intervals for each study site were calculated from weights derived from the PNLO census. Sampling weights were calculated by multiplying the probability of selection at three levels: health area, cluster, and household. Crude and adjusted odds ratios with 95% Confidence Intervals for PCR positivity for *P. falciparum, P. ovale* spp., and *P. vivax* were calculated with sampling weights using multivariate logistic regression with sampling weights in the PROC SURVEYLOGISTIC procedure in SAS 9.4 (SAS Institute Inc., 2024). Multivariate analysis was adjusted for each of the presented covariates. Descriptive statistical analyses and visualizations were done in R version 4.2.2 (R Foundation for Statistical Computing, 2022).

## RESULTS

### Household and participant characteristics

A total of 871 households were visited across 7 health areas with 3,661 participants enrolled from May-September 2023, the latter part of the main rainy season. Patient and household characteristics are described in **Table 3**. Most participants were female (n=2113/3661, 57.7%), and the median age of participants was 22 years old (IQR 13-39), with just over one third of participants under the age of 18 (n=1376/3661, 37.6%). Roughly half (n=1745/3661, 47.7%) of participants reported sleeping under a bednet the night before, and 469 (12.8%) participants had undergone treatment for malaria in the last 30 days. Travel in the 28 days prior to enrollment was uncommon for international destinations with only 13 out of 3661 (0.4%) individuals traveling outside of Cameroon, but 709 (19.4%) participants had recently traveled within the country. The median household size was 6 persons (IQR 4-9). Almost half (n=418/871, 48.0%) of households owned livestock, with chickens (73.9%) and pigs (36.6%) as the most common animals raised.

**Table 3.**
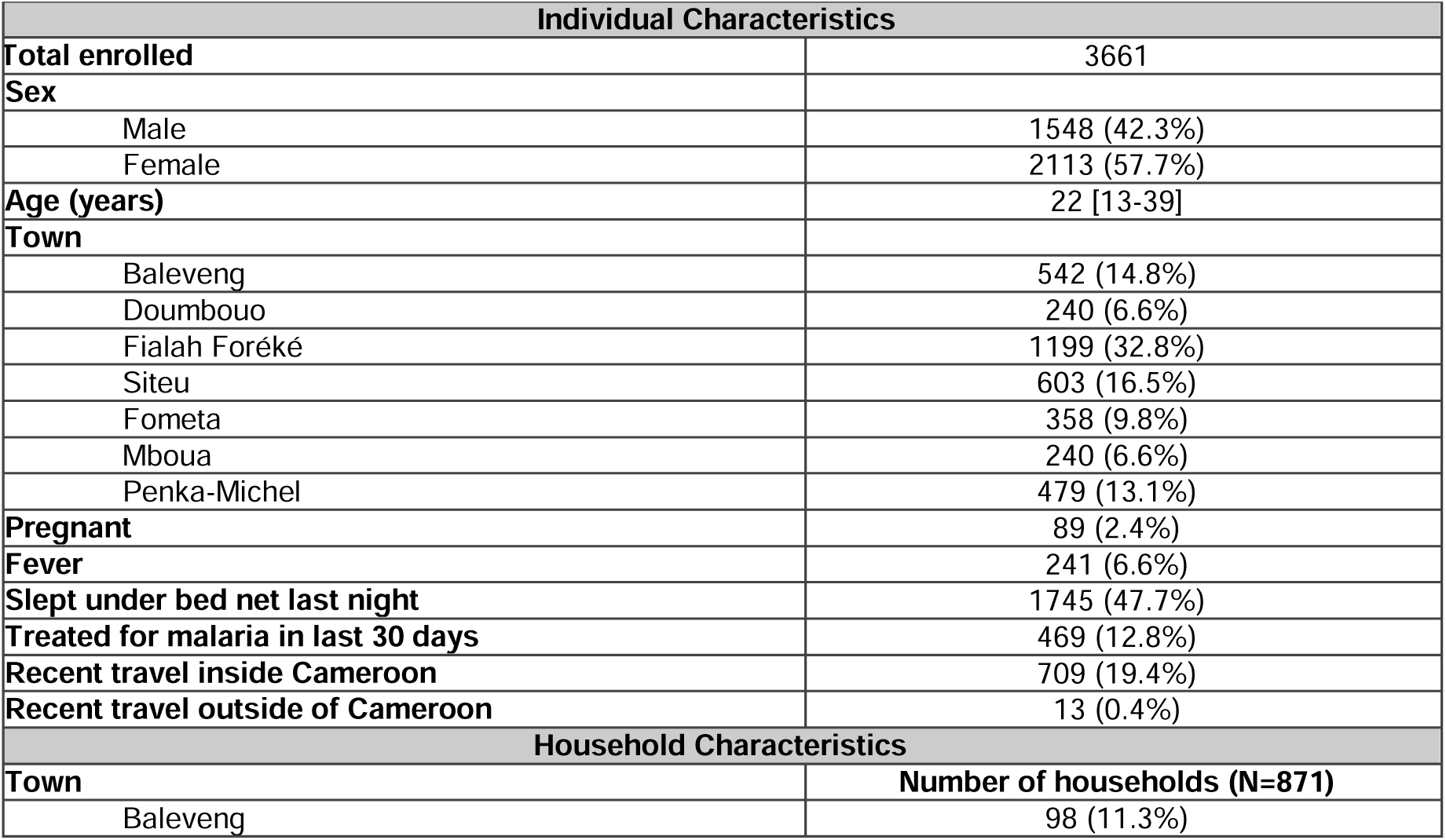
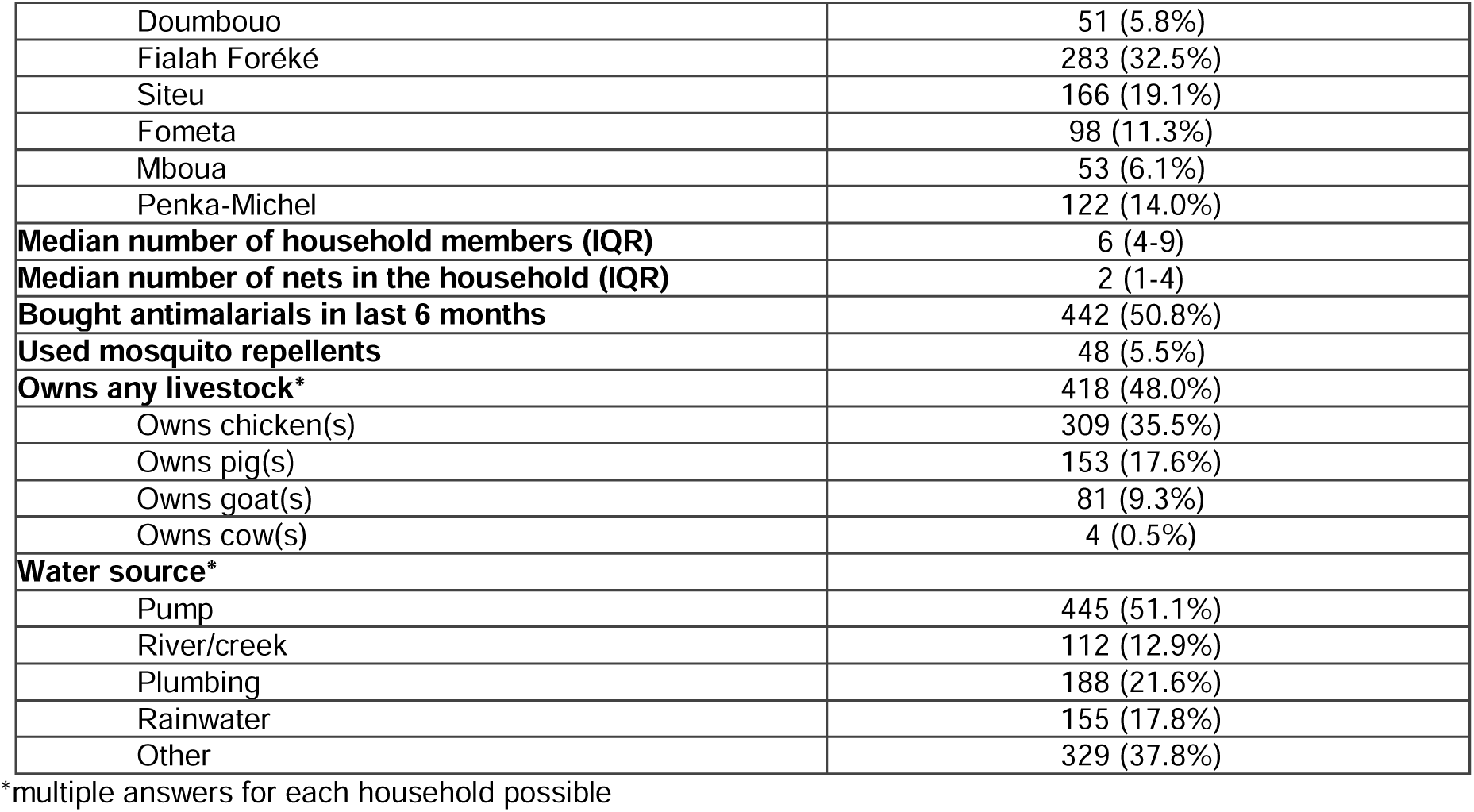
Community survey population and household characteristics.

| Individual Characteristics |  |
| --- | --- |
| Total enrolled | 3661 |
| Sex |  |
| Male | 1548 (42.3%) |
| Female | 2113 (57.7%) |
| Age (years) | 22 [13-39] |
| Town |  |
| Baleveng | 542 (14.8%) |
| Doumbouo | 240 (6.6%) |
| Fialah Foréké | 1199 (32.8%) |
| Siteu | 603 (16.5%) |
| Fometa | 358 (9.8%) |
| Mboua | 240 (6.6%) |
| Penka-Michel | 479 (13.1%) |
| Pregnant | 89 (2.4%) |
| Fever | 241 (6.6%) |
| Slept under bed net last night | 1745 (47.7%) |
| Treated for malaria in last 30 days | 469 (12.8%) |
| Recent travel inside Cameroon | 709 (19.4%) |
| Recent travel outside of Cameroon | 13 (0.4%) |
| Household Characteristics |  |
| Town | Number of households (N=871) |
| Baleveng | 98 (11.3%) |
| Doumbouo | 51 (5.8%) |
| Fialah Foréké | 283 (32.5%) |
| Siteu | 166 (19.1%) |
| Fometa | 98 (11.3%) |
| Mboua | 53 (6.1%) |
| Penka-Michel | 122 (14.0%) |
| **Median number of household members (IQR)** | 6 (4-9) |
| **Median number of nets in the household (IQR)** | 2 (1-4) |
| **Bought antimalarials in last 6 months** | 442 (50.8%) |
| **Used mosquito repellents** | 48 (5.5%) |
| **Owns any livestock\*** | 418 (48.0%) |
| Owns chicken(s) | 309 (35.5%) |
| Owns pig(s) | 153 (17.6%) |
| Owns goat(s) | 81 (9.3%) |
| Owns cow(s) | 4 (0.5%) |
| **Water source\*** |  |
| Pump | 445 (51.1%) |
| River/creek | 112 (12.9%) |
| Plumbing | 188 (21.6%) |
| Rainwater | 155 (17.8%) |
| Other | 329 (37.8%) |

\*multiple answers for each household possible

### Community parasite prevalence

Overall, community carriage of malaria parasites was low when determined by qPCR (**Table 4**). The relapsing malarias were rare across all sites, with only 2 (0.1%; [0%-0.29%]) cases of *P. vivax* and 26 (0.64%; [0.3%-1.0%]) cases of *P. ovale* spp. identified by qPCR. Falciparum malaria was more common with the weighted prevalence of *P. falciparum* at 8.1%; [7.0%-9.2%]. This prevalence was variable across sites with Penka-Michel (19.7%; [15.6%-23.7%), Baleveng (10.0%; [7.4%-12.6%]), and Mboua (8.6%; [4.6%-12.5%]) having the highest weighted prevalence of *P. falciparum.* Both participants with a *P. vivax* infection, and 6/26 (23.1%) of the *P. ovale* spp. cases were coinfections with *P. falciparum.* Fever was reported for 241/3661 participants (weighted proportion 6.3%; 95% CI: [5.2%-7.4%]) with 11.2% (n=27/241) of those with fever positive for *P. falciparum* by RDT. Only 4/26 (15.3%) of the *P. ovale* spp. (including 6 with co-infecting *P. falciparum*) and 36/302 (11.9%) of the *P. falciparum* cases were febrile at the time of sample collection.

**Table 4.**
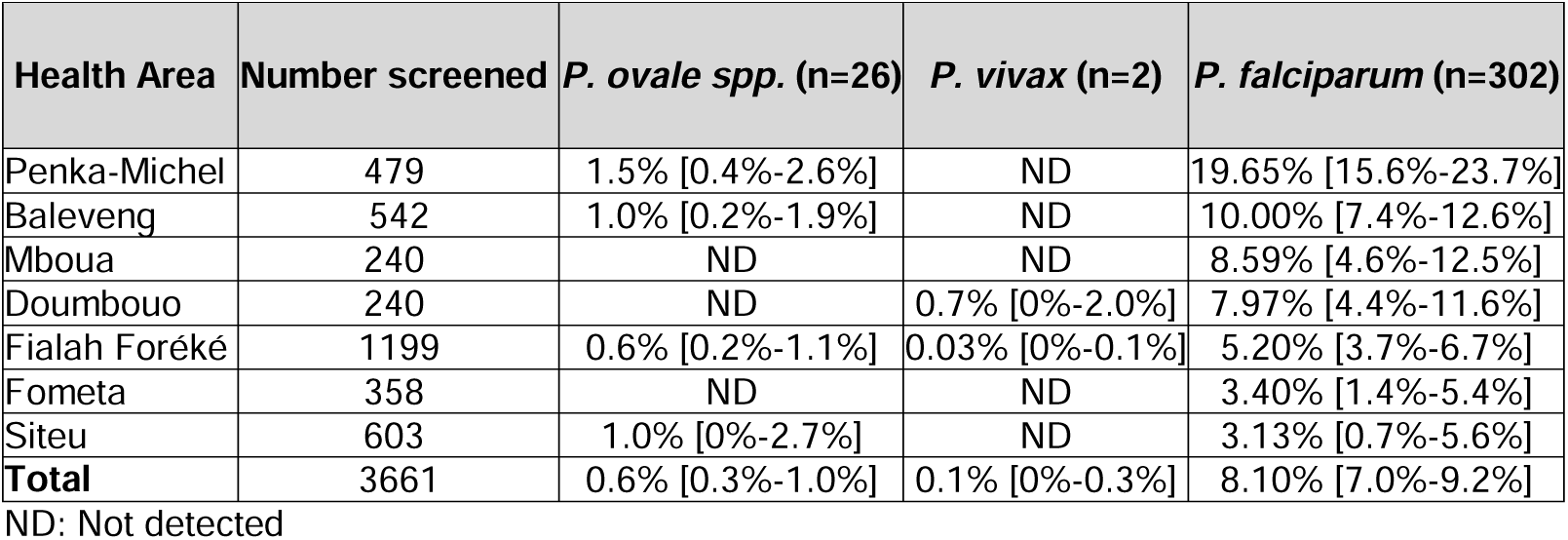
Weighted prevalence and 95% CI of *P. falciparum, P. ovale* and *P. vivax* by site.

Clustering analyses found a statistically significant difference in risk of PCR-positivity for *P. ovale* spp. in Penka-Michel and Baleveng compared to the rest of the study area. Similarly, Penka-Michel, Baleveng, and Mboua had a statistically significant difference in risk of PCR-positivity for *P. falciparum* compared to the rest of the study area (Figure 2).

**Figure 2.**
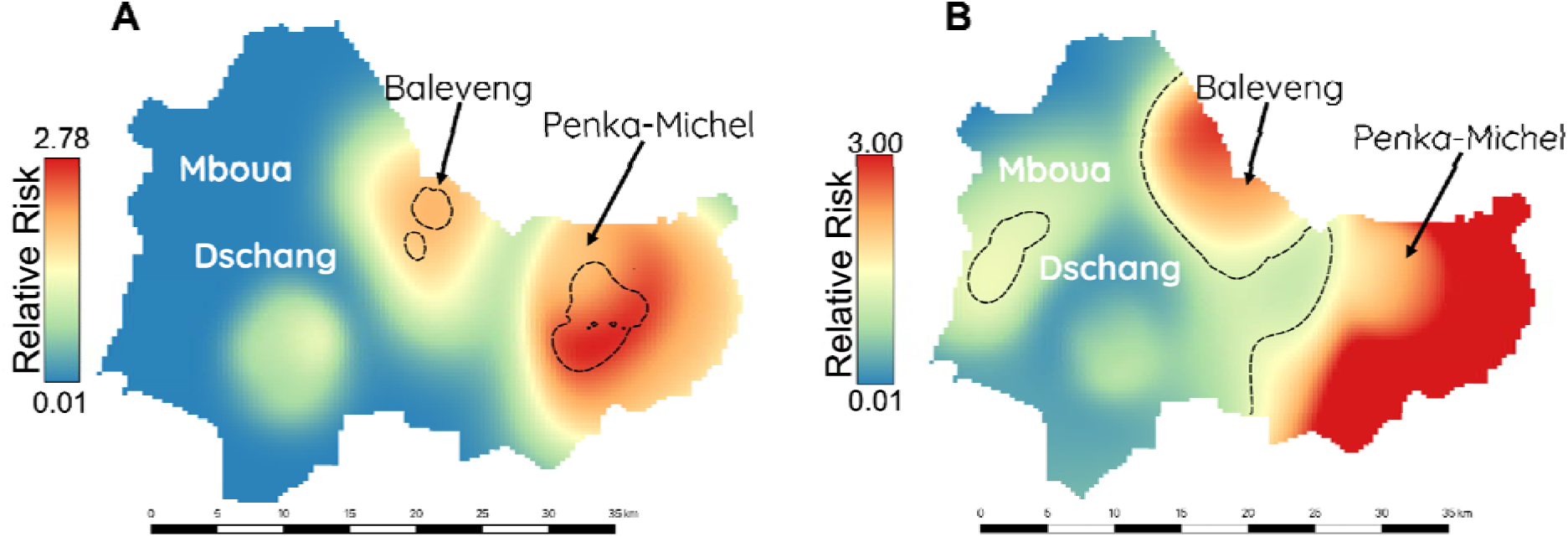
Spatial distribution of (A) *P. ovale* spp. and (B) *P. falciparum*. Black dotted line shows adaptive tolerance contours indicating areas of significantly increased risk of infection.

### Parasite density, gametocytemia and gametocyte assay LOD

Overall, the median parasitemia was 10 (IQR 3.9-64) genome equivalents/µL for *P. ovale* spp. and 16 genome equivalents/µL (IQR: 2.8-138) for *P. falciparum* (**Supplemental Figure 1**). Parasitemia was significantly higher in febrile participants who were PCR-positive for *P. falciparum*, but not for *P. ovale* spp. (**Figure 3**). Gametocytemia was measured in 22/26 *P. ovale* spp. infected individuals. Female gametocytes (*pos25*) for *P. ovale* spp. were detected in 3/22 (14%) samples with a range of 45-228 transcript copies/µL. Male ovale gametocytes (*pomget*) were also detected in 2/22 (9%) samples with 12.0 and 6.4 transcript copies/µL. Gametocytemia was measured in 300/302 samples that were positive for *P. falciparum*. Female gametocytes for *P. falciparum* were detected in 115/300 (38%) with a median gametocytemia of 1.4 gametocytes/µL (IQR: 0.4-5.4 gametocytes/µL). Male gametocytes for *P. falciparum* were detected in 28/300 (9.3%) samples with a median gametocytemia of 2.1 (IQR: 0.5-8.8 gametocytes/µL). There was no correlation between *P. falciparum* female gametocytemia and parasitemia (r = 0.089, **Supplemental Figure 2**). A LOD determined by probit analysis for the *pomget* gametocyte target was estimated at 281 transcript copies per µl (95% CI: 162, 1,310) using 12 replicates across a range of target concentrations. **(Supplemental Figure 3, Supplemental Table 5)**. The interassay coefficient of variation, a measure of precision and repeatability, was determined for the *pomget* assay (**Supplemental Table 6**). Replicates of control plasmids for *pos25*, showed a detection level similar to the previously published LOD, which detected 1 transcript copy per µl in at least 50% of replicates (**Supplementary Table 5**) (46). No false positive results were observed across our non-template controls for all gametocyte detection assays.

**Figure 3.**
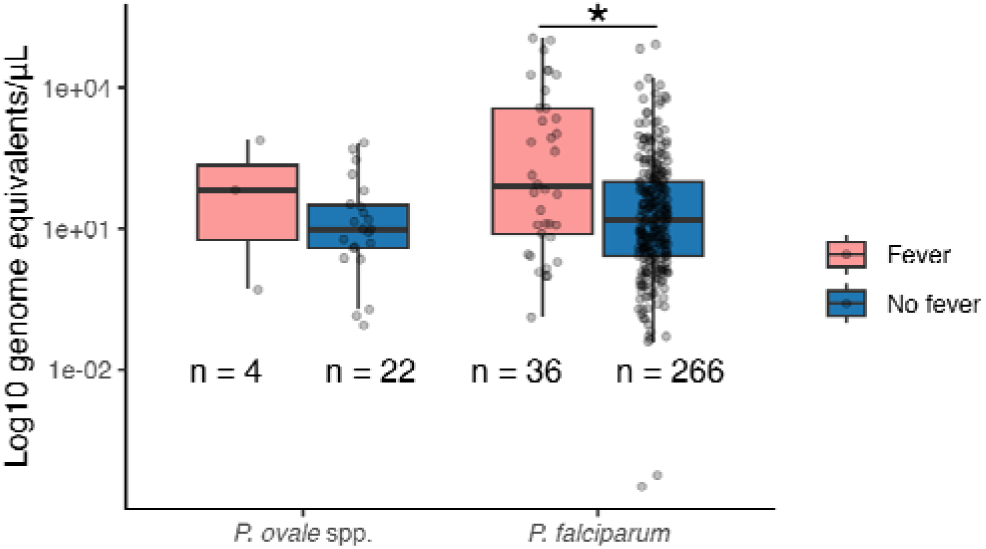
Parasitemia by fever status for *P. falciparum* and *P. ovale* spp. Kruskal-Wallis chi-squared = 9.48, p=0.002 for *P. falciparum* and Kruskal-Wallis chi-squared = 0.45, p-value = 0.50 for *P. ovale* spp. *: significant difference

### Factors associated with *P. ovale* spp. and *P. falciparum* infection

Multivariate logistic regression with sampling weights were used to assess for associations between infection with each species and household and individual characteristics (Figure 4, **Supplemental Table 7**). Children (14 years and younger) had 1.8 (95% CI: [1.3-2.6]) times the adjusted odds of PCR positivity for *P. falciparum* when compared to adults. Participants who had traveled within Cameroon (aOR=2.4; [1.7-3.4]), and those who had received antimalarial treatment in the last 28 days (aOR= 2.1; [1.3-3.4]) had increased adjusted odds of PCR positivity for *P. falciparum*. Over 60% of the travel within Cameroon was to either Bafoussam (159/709, 22%), Douala (149/709, 21%), or Yaoundé (124/709, 18%). No factors associated with an increased odds of PCR positivity for *P. ovale* spp. were identified.

**Figure 4.**
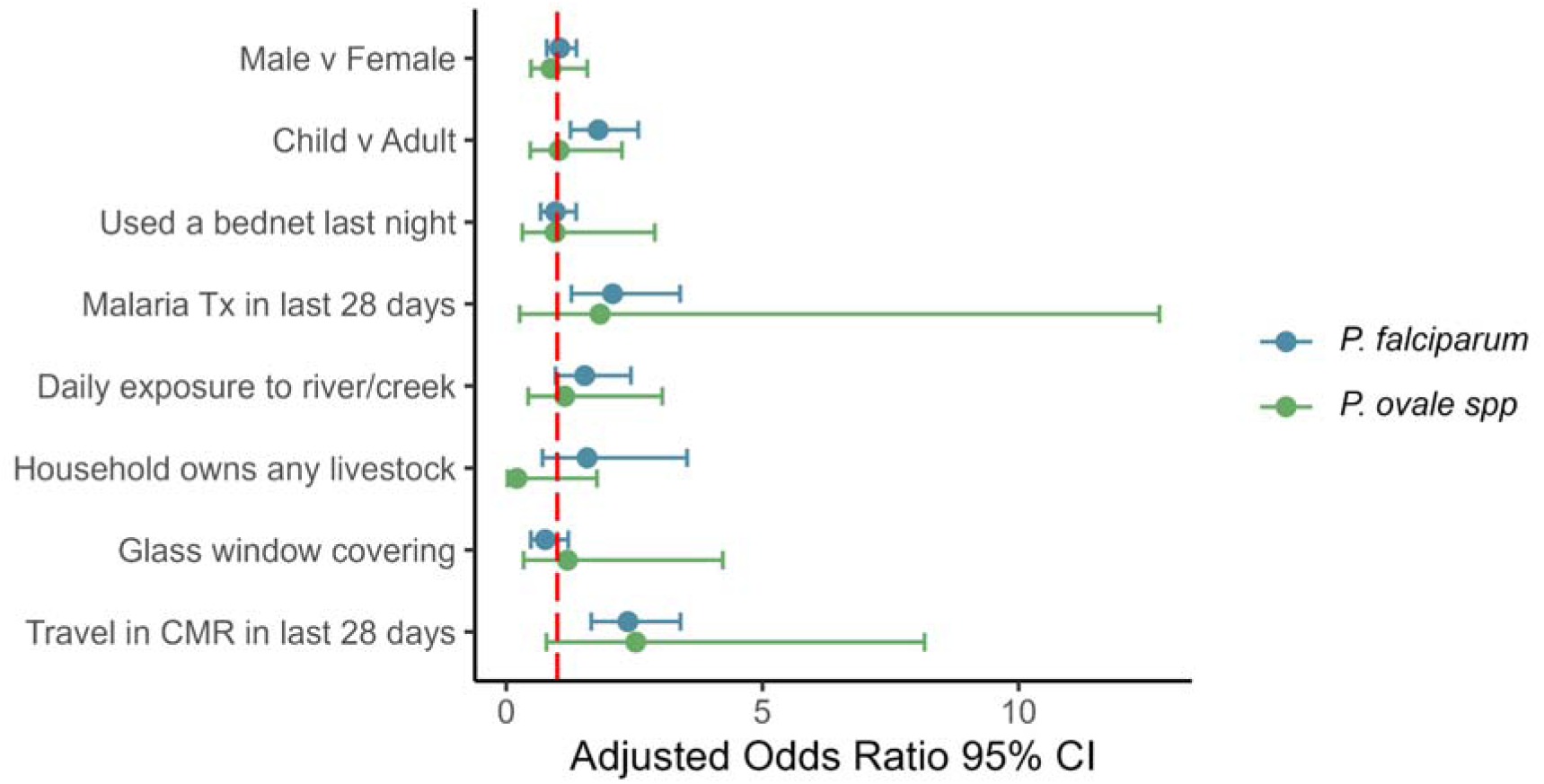
Adjusted odds ratios of infection and 95% confidence intervals for individual and household characteristics. CMR: Cameroon; Tx: treatment

### Mosquito abundance in relation to infection prevalence

We analyzed the subset of mosquito data for areas where human sampling was previously done (54), which provided mosquito density data for Penka-Michel and Dschang, which included study sites Fialah Foréké, Fometa, and Doumbouo (**Figure 1**). Of the 871 households where human sampling occurred, 660 (75.8%) were within 2km of a household where mosquito collections occurred. Due to low mosquito abundance in the Dschang neighborhoods, data from these sites were combined for analysis. Individual neighborhood data in Dschang were similar and are shown in **Supplemental Table 8**. Overall mosquito abundance and human-biting rates (estimated by human landing catches) were higher in Penka-Michel, which had high *P. ovale* spp. and *P. falciparum* infection rates compared to Dschang (**Table 5**). *An. gambiae* (*s.l.*) was the most abundant vector collected at all study sites, accounting for 82.9% (2619/3157) of all mosquitoes captured. Mosquito species composition varied across the sites, with Penka-Michel having significantly greater abundance of all three species identified (*An. gambiae* (*s.l.*)*, An. funestus* (*s.l.*), and *An. ziemanni*) compared to other collection sites (**Table 5** and **Figure 5**). We observed an outdoor shift toward earlier (before 10 PM) human host-seeking behavior for *An. gambiae* (*s.l.*) and a lower abundance of *An. gambiae* (*s.l*.), *An. funestus* (*s.l*.), and *An. ziemanni,* in Dschang compared to Penka-Michel (**Supplemental Table 8** and **Supplemental Figure 4**).

**Figure 5.**
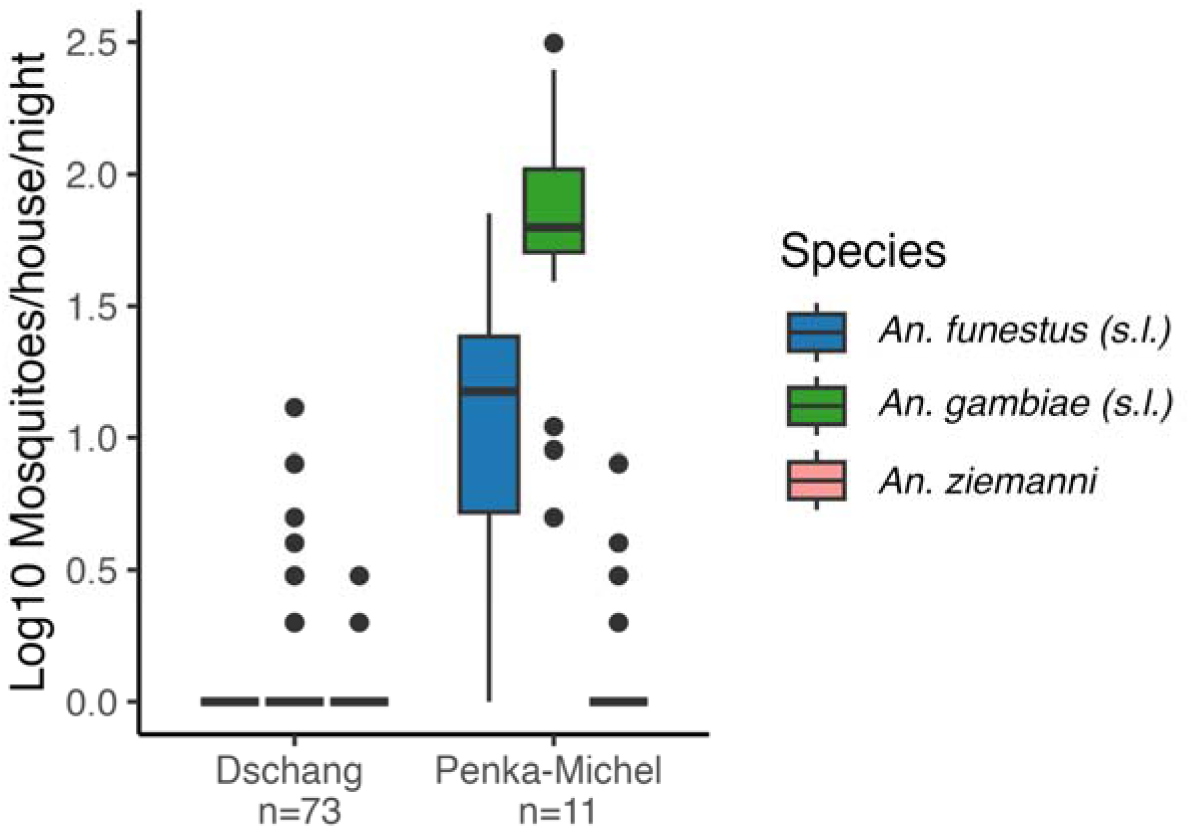
*Anopheles* spp. abundance per night and household by collection.

**Table 5.** Relationship between mosquito abundance, biting rates, and malaria infections.

| Area | Community Po prev | Community Pf prev | Malaria cases/1000 inhabitants<sup>#</sup> | Biting rate (bites/person/night) (54) | *An. gambiae* Median Abundance per Night per House (IQR) | *An. funestus* Median Abundance per Night per House (IQR) | *An. ziemanni* Median Abundance per Night per House (IQR) |
| --- | --- | --- | --- | --- | --- | --- | --- |
| Penka-Michel | 1.5% | 19.7% | 95.5 | 26.2 | 63 (50.5-104.75) | 15 (5.3-24.3) | 0 (0-1) |
| Dschang* | 0.4% | 5.1% | 50.3 | 0.03 | 0 (0-1) | 0 (0-0) | 0 (0-0) |

\*: Dschang represents health zones Fialah Foréké, Fometa and Doumbouo.
#: From national malaria control program data, 2023

## DISCUSSION

This study provides high-quality, community-based molecular survey data of relapsing malaria and falciparum malaria in the Western highlands of Cameroon, centered around Dschang. Overall, relapsing malaria was uncommon in the community. *P. ovale* spp. was found to have an overall prevalence of 0.64% (95% CI: [0.2%-1.0%]), while only two cases of *P. vivax* were detected. *P. falciparum* remained the most common species with an overall prevalence of 8.1% [7.0%-9.2%]. Spatial clustering analysis showed significant differences between health zones, with higher prevalence in Penka-Michel (**Figure 2** and **Table 4**). This was also associated with higher *Anopheles* spp. abundance and higher biting rates (**Table 5** and **Figure 5**). Located roughly 20 kilometers east of Dschang, Penka-Michel is more rural and less densely populated. Baleveng, an outlying village near Dschang, showed the second highest prevalence for both *P. ovale* spp. and *P. falciparum*, with statistically significant clustering (**Figure 2**). Mboua showed an increased risk of *P. falciparum*; however, no cases of *P. ovale* spp. were detected in Mboua.

Our results are different from previous studies of relapsing malaria that have focused on clinic populations in Dschang. Russo et al found *P. vivax* infection in 5.6% of febrile outpatients, while Djeunang Dongho *et al*. reported that 35.2% of febrile patients in the hospital were PCR-positive for *P. vivax* during the rainy season (14,31). This high rate of vivax positivity in the clinic is very distinct from the negligible number of cases we identified in this predominantly asymptomatic community survey. Based on these previous studies, the near absence of *P. vivax* from the study population was unexpected. Potential explanations for the lack of *P. vivax* cases in our study population include the resolution of a previous outbreak captured in previous studies, overall low malaria transmission during the study period, inaccessible sources of previous *vivax* outbreaks like short-term economic migrants to Dschang, or the initial reports represented initial outbreaks, which resulted in population immunity. These possibilities would need to be assessed by different studies.

As this was a community survey, most infections were asymptomatic, with only 6.6% of participants febrile at the time of sample collection. A substantial proportion of ovale infections (23%) were mixed with *P. falciparum*. In general, low case counts for *P. ovale* resulted in wide confidence intervals in risk factor assessment and limited our ability to find significant associations. Despite other evidence that school-aged children (6-18 years old) act as a significant asymptomatic reservoir for non-falciparum malaria (55), we did not find increased odds of PCR positivity for *P. ovale* in this age group compared to adults. This was true for both mono-species *P. ovale* or for all *P. ovale* infections. This is consistent with findings in Tanzania, where risk of infection was not different for school-aged children compared to adults (21). However for *P. falciparum*, being school-aged was a significant predictor of PCR positivity (aOR = 1.6; [1.2-2.2]) consistent with previous studies (56,57). Recent travel within Cameroon was also associated with over twice the odds of PCR positivity for *P. falciparum*, which may indicate that these cases were contracted while traveling to or through higher transmission areas. Three destinations accounted for over 60% of travel within Cameroon: Bafoussam (n=159/709), Yaoundé (n=124/709), and Douala (n=149/709). Bafoussam is the capital and economic center for the West Region of Cameroon and is located approximately 50 km from Dschang with an estimated population of 480,000 (58). It is known to be endemic for malaria, with the vast majority being *P. falciparum*, but *P. vivax* has been detected (28,59). Yaoundé is the capital of Cameroon, with a population exceeding 4.5 million people, and is located 238 km away from Dschang. Malaria remains common in the urban and peri-urban areas of Yaoundé, with asymptomatic carriage rates in children reaching over 70% (60). Asymptomatic carriage in adults is also common in the surrounding areas of Yaoundé, with 81% of *P. falciparum* positive adults in a longitudinal household without symptoms (61). Douala is the major port and economic capital for Cameroon, with a population of over 3.5 million people, and is 159 km from Dschang. *P. falciparum* infections are common and *P. ovale* has also been reported in this region (62,63).

A previous study of *P. ovale* spp. gametocytemia in Papua New Guinea reported a low gametocyte carriage rate (2.2%, 11/505) in the population, with 64.7% of *P. ovale* infected individuals (11/17) being gametocytemic (46). This study leveraged a female-specific assay targeting the orthologue of *pfs25* with a LOD estimated at 1 copy/μL (46). We found a significantly lower rate of detectable gametocytes [8.7% (3/22)] among *P. ovale* spp. qPCR-positive samples using this assay despite a similar LOD. Our new *P. ovale* spp. male-specific assay (*pomget*) detected the same proportion of gametocytemia as the *pos25* assay [14% (3/22)], but with a higher assay LOD. While sample quality and RNA extraction efficiency could cause underdetection, *P. falciparum* gametocytemia was relatively high in our cohort, with 38% (115/302) of qPCR-positive samples also having detectable levels of gametocytes. This rate is higher to a previous study in the Center Region of Cameroon that found a gametocyte positivity rate of 10% (32/317) using the *pfs25* assay among individuals who were infected with *P. falciparum,* as determined by RDT or LAMP (64). It is intriguing that such low gametocytemia rates were detected given human skin feeding studies in Tanzania, which did not quantify *P. ovale* spp. gametocytemia burden, have shown high infectivity (65). It suggests that there may be undetermined biological factors (highly competent malaria vectors, contribution of hypnozoite-induced relapses, frequent but transient parasitemia/gametocytemia) that help support community transmission of *P. ovale* spp.

As expected, malaria rates were higher in areas with greater *Anopheles* vector abundance and elevated biting rates (**Table 5**). Consistent with this, previous analyses revealed that mosquito infection rates were higher in Penka-Michel compared to Dschang, with the majority of infected mosquitoes in Penka-Michel carrying *P. falciparum* (54). Penka-Michel is a more rural, less populated and more densely vegetated area compared to Dschang. In Penka-Michel, the overall infection rate was 3.2% (13/392), and three mosquito species tested positive for malaria: *Anopheles gambiae*, *An. funestus*, and *An. ziemanni* (54). Among the infected mosquitoes, *P. ovale* was detected in 7.7% (1/13), identified in a *An. gambiae* (*s.l.*) mosquito (54). Unfortunately, mosquito abundance in Dschang was too low to support meaningful analyses of associations with environmental variables (**Tables 2** and **3**).

While this study provides the most detailed study of relapsing and falciparum malaria in the communities around Dschang in the Western Highlands, there are several notable limitations. First, our sampling was confined to late in the malaria transmission season. Reports from Tanzania have suggested that *P. ovale* spp. infection peaks early in the transmission season (65). Second, while the associations between malaria prevalence, mosquito abundance, and biting rates are as expected, the geographic overlap of sampling of households and vectors is not one-to-one,with only 75.8% of human sampling households within 2km of a mosquito collection site. In addition, mosquito collection did not occur in two sites: Mboua had inadequate mosquito abundance, and collections in Baleveng are planned for a future study. This may introduce bias into these associations as mosquito exposure in the human sampling population is only approximated by the mosquito collections done. Lastly, children younger than 6 years of age were not sampled, excluding an age group expected to have lower immunity and higher risk of adverse health outcomes associated with exposure.

## CONCLUSION

This is the first community molecular survey of falciparum and the relapsing malarias in and around Dschang, Cameroon, an area with previous evidence of waning *P. falciparum* predominance in clinic populations (14,24,27,31). Overall, prevalence of relapsing malaria was low among the 3,661 participants. Malaria was heterogeneously distributed across sampled communities, with prevalence ranging from 19.7% to 3.1% for *P. falciparum* and 1.5% to 0% for *P. ovale* spp.,and as expected, vector abundance was associated with higher malaria rates within the community. These data are at odds with previous reports of *P. vivax* infection being common among clinic patients in this community and raises questions about the dynamics of non-falciparum malaria within the region and its burden in the community. Furthermore, gametocytemia was low among those infected with *P. ovale*, in contrast to *P. falciparum* infections. The Relapsing Malaria in Africa (ReMA) study is continuing to gather data from both community surveys like the one described herein, as well as clinic based surveillance and paired vector surveillance, to better understand the potentially dynamic epidemiology of relapsing malaria in the region.

## Data Availability

Metadata for this study is available at UNCs Dataverse at doi: https://doi.org/10.15139/S3/JQXDZ0, except for geographic location which is available from Dr. Ali upon reasonable request. Mosquito data is available from Kiam et al. 2025 on request, doi: https://doi.org/10.21203/rs.3.rs-5558659/v1.

https://doi.org/10.21203/rs.3.rs-5558659/v1

## Acknowledgements

We thank the participants of the study. We thank the members of the ReMA team for the work that went into conducting the community survey. We thank Oksana Kharabora and Catherine Gorman for assistance in the lab. The following reagent was obtained through BEI Resources, NIAID, NIH: Diagnostic Plasmid Containing the Small Subunit Ribosomal RNA Gene (18S) from *Plasmodium falciparum*, MRA-177, contributed by Peter A. Zimmerman. The following reagent was obtained through BEI Resources, NIAID, NIH: Diagnostic Plasmid Containing the Small Subunit Ribosomal RNA Gene (18S) from *Plasmodium vivax*, MRA-178, contributed by Peter A. Zimmerman. The following reagent was obtained through BEI Resources, NIAID, NIH: Diagnostic Plasmid Containing the Small Subunit Ribosomal RNA Gene (18S) from *Plasmodium ovale*, MRA-180, contributed by Peter A. Zimmerman

## Author Contributions

**Data Curation:** SJW, VPKT, MM, AGBT and BK

**Formal Analysis:** SJW, VPKT, and VG

**Funding Acquisition:** JJJ, RRD, IMA and SEN

**Investigation:** VPKT, MM, CG, LKM, ZPH, KCE, EH, JMS, AGBT, BK, MNN, DBK, GNL, CLD, CAN, MN, II, JCO FCL and YNN

**Methodology:** JJJ, SEN, RRD, JTL, JAB and IMA.

**Project Administration:** RRR, SEN, RRD, JTL and IMA.

**Resources:** JJJ, RRD, IMA and SEN

**Software:** SJW and VG

**Supervision:** RRR, SEN, RRD, JTL and IMA.

**Validation:** SJW, VG and VPKT

**Visualization:** SJW and VG

**Writing – Original Draft Preparation:** SJW, JJJ, JTL

**Writing – Review & Editing:** All authors

## Data Availability

Metadata for this study is available at UNC’s Dataverse at doi: https://doi.org/10.15139/S3/JQXDZ0, except for geographic location which is available from Dr. Ali upon reasonable request. Mosquito data is available from Kiam et al. 2025 on request, doi: https://doi.org/10.21203/rs.3.rs-5558659/v1.

