## Supplemental Material for "Epidemiology of Relapsing and Falciparum Malaria in the Highlands of Cameroon: An Integrated Community Survey of Human Infection and Vector Abundance"

Supplemental Table 1…………………………………………………………………….. 2
Supplemental Table 2…………………………………………………………………….. 3
Supplemental Table 3…………………………………………………………………….. 4
Supplemental Table 4…………………………………………………………………….. 5
Supplemental Table 5…………………………………………………………………….. 6
Supplemental Table 6…………………………………………………………………….. 7
Supplemental Table 7…………………………………………………………………….. 8
Supplemental Table 8…………………………………………………………………….. 9

**Supplemental Table 1. Sampling Weights**

| Health Area | Village | Number of Clusters | Probability of Cluster Selected | Estimated Number of Households | Number of Households Selected | Probability of Household Selected | Weight |
|---|---|---|---|---|---|---|---|
| Baleveng | Menka | 21 | 0.286 | 291 | 15 | 0.05 | 67.85 |
| Baleveng | Lepang | 21 | 0.286 | 282 | 10 | 0.04 | 98.84 |
| Baleveng | Dijo | 21 | 0.286 | 491 | 20 | 0.04 | 85.89 |
| Baleveng | Zem | 21 | 0.286 | 204 | 24 | 0.12 | 29.75 |
| Baleveng | Nza'ah | 21 | 0.286 | 565 | 28 | 0.05 | 70.68 |
| Baleveng | Melio | 21 | 0.286 | 180 | 1 | 0.01 | 628.60 |
| Doumbouo | Suella Batsingla | 20 | 0.2 | 280 | 13 | 0.05 | 107.69 |
| Doumbouo | Kamnick | 20 | 0.2 | 126 | 12 | 0.10 | 52.42 |
| Doumbouo | Tsing Saah | 20 | 0.2 | 334 | 12 | 0.04 | 139.08 |
| Doumbouo | Saah | 20 | 0.2 | 150 | 14 | 0.09 | 53.57 |
| Fialah Foréké | Zembing | 21 | 0.667 | 137 | 30 | 0.22 | 6.85 |
| Fialah Foréké | Mingmeto | 21 | 0.667 | 384 | 38 | 0.10 | 15.16 |
| Fialah Foréké | Femlah | 21 | 0.667 | 266 | 13 | 0.05 | 30.69 |
| Fialah Foréké | Mingou route | 21 | 0.667 | 454 | 10 | 0.02 | 68.10 |
| Fialah Foréké | Ngui 3 | 21 | 0.667 | 205 | 13 | 0.06 | 23.65 |
| Fialah Foréké | Vallee | 21 | 0.667 | 368 | 29 | 0.08 | 19.03 |
| Fialah Foréké | Lefock | 21 | 0.667 | 258 | 27 | 0.10 | 14.33 |
| Fialah Foréké | Ngui 1 | 21 | 0.667 | 488 | 25 | 0.05 | 29.28 |
| Fialah Foréké | Doungha | 21 | 0.667 | 548 | 18 | 0.03 | 45.67 |
| Fialah Foréké | Meka'a | 21 | 0.667 | 819 | 30 | 0.04 | 40.95 |
| Fialah Foréké | Toumeto | 21 | 0.667 | 305 | 12 | 0.04 | 38.13 |
| Fialah Foréké | Mechieu | 21 | 0.667 | 437 | 10 | 0.02 | 65.55 |
| Fialah Foréké | Ngui 2 | 21 | 0.667 | 344 | 12 | 0.03 | 43.00 |
| Fialah Foréké | King place | 21 | 0.667 | 270 | 16 | 0.06 | 25.31 |
| Fometa | Fiakop 2 | 16 | 0.375 | 205 | 22 | 0.11 | 24.85 |
| Fometa | Keleng 1 | 16 | 0.375 | 251 | 14 | 0.06 | 47.81 |
| Fometa | Touleppe | 16 | 0.375 | 195 | 14 | 0.07 | 37.14 |
| Fometa | Lefatsa | 16 | 0.375 | 204 | 15 | 0.07 | 36.27 |
| Fometa | Fialah | 16 | 0.375 | 289 | 17 | 0.06 | 45.33 |
| Fometa | Minpeuh | 16 | 0.375 | 159 | 16 | 0.10 | 26.50 |
| Mboua | Mboua | 15 | 0.267 | 303 | 16 | 0.05 | 71.02 |
| Mboua | Apang | 15 | 0.267 | 58 | 10 | 0.17 | 21.75 |
| Mboua | Dzifoda | 15 | 0.267 | 128 | 14 | 0.11 | 34.29 |
| Mboua | Fossong | 15 | 0.267 | 352 | 13 | 0.04 | 101.54 |
| Penka-Michel | Bagham | 19 | 0.421 | 116 | 13 | 0.11 | 21.19 |
| Penka-Michel | lac | 19 | 0.421 | 121 | 15 | 0.12 | 19.16 |
| Penka-Michel | lonako | 19 | 0.421 | 319 | 15 | 0.05 | 50.51 |
| Penka-Michel | Nylon | 19 | 0.421 | 178 | 18 | 0.10 | 23.49 |
| Penka-Michel | Plateau | 19 | 0.421 | 247 | 15 | 0.06 | 39.11 |
| Penka-Michel | Tami | 19 | 0.421 | 96 | 17 | 0.18 | 13.41 |
| Penka-Michel | Tergal | 19 | 0.421 | 130 | 14 | 0.11 | 22.05 |
| Penka-Michel | Hopital | 19 | 0.421 | 89 | 15 | 0.17 | 14.09 |
| Siteu | Tapale | 15 | 0.667 | 171 | 16 | 0.09 | 16.03 |
| Siteu | Nylon | 15 | 0.667 | 151 | 15 | 0.10 | 15.10 |
| Siteu | Tsinkop | 15 | 0.667 | 242 | 23 | 0.10 | 15.78 |
| Siteu | Tchouale 2 | 15 | 0.667 | 242 | 17 | 0.07 | 21.35 |
| Siteu | Madagascar | 15 | 0.667 | 1700 | 15 | 0.01 | 169.92 |
| Siteu | Tchouale 1 | 15 | 0.667 | 292 | 19 | 0.07 | 23.04 |
| Siteu | Le tchouala | 15 | 0.667 | 183 | 17 | 0.09 | 16.15 |
| Siteu | Tsinfem | 15 | 0.667 | 174 | 17 | 0.10 | 15.35 |
| Siteu | Haoussa | 15 | 0.667 | 143 | 15 | 0.10 | 14.30 |
| Siteu | Lefang | 15 | 0.667 | 86 | 12 | 0.14 | 10.75 |

**Supplemental Table 2. Malaria qPCR Detection Methods**

| | | | | |
|---|---|---|---|---|
| ***Plasmodium falciparum (18s)*** | | | | |
| **Adapted from:** | Veron V et al. Exp Parasitol 2009. 121(4):346-51. | | | |
| **Forward Primer** | 5' - ATTGCTTTTGAGAGGTTTTGTTACTTT - 3' | | | |
| **Reverse Primer** | 5' - GCTGTAGTATTCAAACACAATGAACTCAA - 3' | | | |
| **Probe** | 5' - FAM/ CATAACAGACGGGTAGTCAT /NFQ - 3' | | | |
| **Cycling conditions** | 1 cycle | 1 cycle | 45 cycles | |
| **Temperature (℃)** | 50 | 95 | 95 | 60 |
| **Time** | 2 min | 10 min | 15 s | 1 min |
| **Reaction conditions:** | PerfeCTa qPCR Tough Mix (QuantaBio, Beverly, MA, USA) | | | |
| | Fwd primer | 300 nM | | |
| | Rev primer | 300 nM | | |
| | Probe | 200 nM | | |
| | Template DNA | 2 µl | | |
| | Total volume | 12 µl | | |
| ***Plasmodium ovale spp.* (18s)** | | | | |
| **Adapted from:** | Mitchell C, et. al. Journal of Infectious Diseases. 2021 | | | |
| **Forward Primer** | 5' - CCRACTAGGTTTTGGATGAAAVRTTTTT- 3' | | | |
| **Reverse Primer** | 5' - AACCCAAAGACTTTGATTTCTCATAA - 3' | | | |
| **Probe** | 5' - VIC/CRA AAGGAATTYTCTTATT /MGB- 3' | | | |
| **Cycling conditions** | 1 cycle | 1 cycle | 45 cycles | |
| **Temp(℃)** | 50 | 95 | 95 | 52 |
| **Time** | 2 min | 10 min | 15 s | 1 min |
| **Reaction:** | PerfeCTa qPCR Tough Mix (QuantaBio, Beverly, MA, USA) | | | |
| | Fwd primer | 400 nM | | |
| | Rev primer | 400 nM | | |
| | Probe | 200 nM | | |
| | Template DNA | 2 µl | | |
| | Total volume | 12 µl | | |
| ***Plasmodium vivax* (18s)** | | | | |
| **Adapted from:** | Brazeau N, et. al. Nature Communications. 2021 | | | |
| **Forward Primer** | 5' - ACGCTTCTAGCTTAATCCACATAACT - 3' | | | |
| **Reverse rimer** | 5' - ATTTACTCAAAGTAACAAGGACTTCCAAGC - 3' | | | |
| **Probe** | 5' - /56-FAM/TTCGTATCG/ZEN/ACTTTGTGCGCATTTTGC/3IABkFQ/ - 3' | | | |
| **Cycling conditions** | 1 cycle | 1 cycle | 45 cycles | |
| **Temperature (℃)** | 50 | 95 | 95 | 60 |
| **Time** | 2 min | 10 min | 15 s | 1 min |
| **Reaction:** | PerfeCTa qPCR Tough Mix (QuantaBio, Beverly, MA, USA) | | | |
| | Fwd primer | 400 nM | | |
| | Rev primer | 400 nM | | |
| | Probe | 200 nM | | |
| | Template DNA | 5 µl | | |
| | Total volume | 18 µl | | |

**Supplemental Table 3.** ***P. falciparum*** **Gametocyte Detection by qPCR Methods**

| | | | | |
|---|---|---|---|---|
| ***P. falciparum*** **male (*pfmget*)** | | | | |
| **Forward Primer** | 5' – GGTCCAAATATAAAATCCTGTTC-3' | | | |
| **Reverse Primer** | 5' – TGTGTAACGTATGATTCATTTTC-3' | | | |
| **Probe** | 5' – FAM -5`CAGCTCCAGCATTAAAAACAC-BHQ1- 3' | | | |
| ***P. falciparum*** **female (*pfs25*)** | | | | |
| **Forward Primer** | GAA ATC CCG TTT CAT ACG CTT G | | | |
| **Reverse Primer** | AGT TTT AAC AGG ATT GCT TGT ATC TAA | | | |
| **Probe** | [AminoC6+HEX]-TGT AAG AAT GTA ACT TGT GGT AAC GGT-[BHQ1a~Q] | | | |
| **Duplexed *P. falciparum*** | | | | |
| **Cycling conditions** | 1 cycle | 1 cycle | 45 cycles | |
| **Temperature (℃)** | 50 | 95 | 95 | 59 |
| **Time** | 15 min | 2 min | 15 sec | 1 min |
| **Reagent** | **Volume (μL)** | | | |
| **SuperScript III RT Platinum Taq Mix** | 0.5 | | | |
| **2X Reaction Mix with ROX** | 12.5 | | | |
| **pfs25 fwd primer (10 μM)** | 2 | | | |
| **pfs25 rev primer (10 μM)** | 2 | | | |
| **pfs25 probe (10 μM)** | 0.2 | | | |
| **pfmget fwd primer (10 μM)** | 2 | | | |
| **pfmget rev primer (10 μM)** | 2 | | | |
| **pfmget probe (10 μM)** | 0.2 | | | |
| **H2O** | 0.6 | | | |
| **Template RNA** | 3 | | | |
| **Total volume/reaction** | 25 | | | |

**Supplemental Table 4.** ***P. ovale spp*** **Gametocyte Detection by qPCR Methods**

| | | | | |
|---|---|---|---|---|
| ***P. ovale mget*** **(PocGH01_12044900_p1 & PowCR01_120040400_p1)** | | | | |
| **Pf ortholog** | pfmGET | | | |
| **Forward Primer (P. ovale curtisi)** | CTTCGCATCCCCAGATTTCC | | | |
| **Reverse Primer (P. ovale curtisi)** | GCCATGTTTGCTTAATTGCCC | | | |
| **Probe** | FAM-AAAAGAAGC/ZEN/AAAGAACTCAAAGG-/ 3IABkFQ/ | | | |
| **Forward Primer (P. ovale walikeri)** | CGCATACCCAGAATTACCCC | | | |
| **Reverse Primer (P. ovale walikeri)** | GTCTCCTTCTCCTGCCTGAG | | | |
| **Cycling conditions** | 1 cycle | 1 cycle | 45 cycles | |
| **Temperature (℃)** | 50 | 95 | 95 | 55 |
| **Time** | 15 min | 2 min | 15 sec | 30 sec |
| **Reagent** | **Volume (uL)** | | | |
| **SS3 RT Platinum Taq Mix** | 0.5 | | | |
| **2X Reaction Mix with ROX** | 12.5 | | | |
| **poc_male forward (10 μM)** | 2 | | | |
| **poc_male reverse (10 μM)** | 2 | | | |
| **po_male probe (10 μM)** | 0.4 | | | |
| **pow_male forward (10 μM)** | 2 | | | |
| **pow_male reverse (10 μM)** | 2 | | | |
| **H2O** | 0.6 | | | |
| **Template RNA/plasmid** | 3 | | | |
| **Total volume/reaction (μL)** | 25 | | | |
| ***pos25*** | | | | |
| **Forward Primer** | CGTACCCGCTGAATGCAAAG | | | |
| **Reverse Primer** | GCCTATATTACATGAGCATCTACC | | | |
| **Probe** | FAM-AACCCAAGCCCGGATAAT–MGB –EQ | | | |
| **Cycling conditions** | 1 cycle | 1 cycle | 45 cycles | |
| **Temperature (℃)** | 55 | 95 | 95 | 58 |
| **Time** | 15 min | 1 in | 10 sec | 1 min |
| **Reagent** | **Volume (μL)** | | | |
| **SS3 RT Platinum Taq Mix** | 0.5 | | | |
| **2X Reaction Mix with ROX** | 12.5 | | | |
| **Pos25 forward (10 μM)** | 2 | | | |
| **Pos25 reverse (10 μM)** | 2 | | | |
| **Pos25 probe (10 μM)** | 0.6 | | | |
| **H2O** | 4.4 | | | |
| **Template RNA/plasmid** | 3 | | | |
| **Total volume/reaction (μL)** | 25 | | | |

**Supplemental Table 5. Limit of detection for *P. ovale spp.* gametocyte assay**

| ***pomget*** | | |
|---|---|---|
| **Copies/$\mu$L** | **Replicates amplified** | **Replicates run** |
| 2871 | 12 | 12 |
| 236 | 11 | 12 |
| 47 | 2 | 12 |
| 9 | 0 | 12 |
| 2 | 0 | 12 |
| ***pos25*** | | |
| **Copies/$\mu$L** | **Replicates amplified** | **Replicates run** |
| 200800 | 5 | 5 |
| 40160 | 5 | 5 |
| 8032 | 5 | 5 |
| 1606.4 | 5 | 5 |
| 321.3 | 5 | 5 |
| 64.3 | 5 | 5 |
| 12.9 | 4 | 5 |
| 2.6 | 3 | 5 |
| 0.5 | 0 | 5 |

**Supplemental Table 6. Interassay and intra-assay coefficient of variation (CV) for male *P. ovale* gametocyte detection using *pomget*. Three serially diluted standards in triplicate across 5 different qPCR runs.**

| Intra-assay CV of *pomget* | | | |
|---|---|---|---|
| **Transcript copies/μL** | **Mean %CV of Ct** | **%CV IQR** | |
| 287100 | 1.45% | 0.96% - 2.05% | |
| 28710 | 2.07% | 0.69% - 2.81% | |
| 2871 | 1.36% | 0.50% - 2.27% | |
| **Interassay CV** | | | |
| **Transcript copies/μL** | **Mean of means (Ct)** | **SD** | **%CV** |
| 287100 | 23.87 | 1.38 | 5.80% |
| 28710 | 29.82 | 1.45 | 4.87% |
| 2871 | 33.46 | 1.84 | 5.48% |

**Supplemental Table 7. Weighted bivariate and adjusted odds ratios with 95% CI**

| ***P. falciparum*** | **OR (95% CI)** | **p-value** | **aOR (95% CI)** | **p-value** |
|---|---|---|---|---|
| Male v Female | 1.15 (0.85-1.57) | 0.36 | 1.04 (0.8-1.37) | 0.75 |
| Child v Adult | 1.53 (1.12-2.08) | 0.007 | 1.80 (1.25-2.58) | 0.002 |
| Used a bednet last night | 0.90 (0.67-1.23) | 0.51 | 0.96 (0.67-1.37) | 0.82 |
| Malaria Tx in last 28 days | 2.05 (1.38-3.04) | 0.0004 | 2.08 (1.27-3.39) | 0.004 |
| Daily exposure to river/creek | 1.57 (1.16-2.12) | 0.004 | 1.53 (0.97-2.44) | 0.07 |
| Household owns any livestock | 1.32 (0.95-1.83) | 0.098 | 1.58 (0.71-3.53) | 0.26 |
| Owns chickens | 1.01 (0.75-1.37) | 0.94 | 0.73 (0.32-1.64) | 0.43 |
| Owns cows | 0.16 (0.02-1.2) | 0.074 | 0.11 (0.01-1.41) | 0.09 |
| Owns goats | 1.05 (0.72-1.54) | 0.78 | | |
| Owns pigs | 1.02 (0.71-1.46) | 0.91 | 0.89 (0.51-1.56) | 0.67 |
| Glass window covering | 0.70 (0.5-0.97) | 0.034 | 0.76 (0.48-1.21) | 0.24 |
| Travel in CMR in last 28 days | 2.03 (1.44-2.86) | 0.0001 | 2.38 (1.66-3.4) | <.0001 |
| Travel outside CMR in last 28 days | 2.01 (0.25-15.98) | 0.51 | 2.44 (0.42-14.06) | 0.31 |
| ***P. ovale spp*** | **OR (95% CI)** | **p-value** | **aOR (95% CI)** | **p-value** |
| Male v Female | 0.92 (0.32-2.66) | 0.88 | 0.87 (0.48-1.58) | 0.65 |
| Child v Adult | 0.81 (0.27-2.42) | 0.70 | 1.03 (0.47-2.26) | 0.94 |
| Used a bednet last night | 0.94 (0.3-2.99) | 0.91 | 0.95 (0.31-2.9) | 0.93 |
| Malaria Tx in last 28 days | 2.00 (0.34-11.72) | 0.44 | 1.83 (0.26-12.74) | 0.53 |
| Daily exposure to river/creek | 1.05 (0.35-3.17) | 0.93 | 1.14 (0.43-3.05) | 0.78 |
| Household owns any livestock | 0.73 (0.22-2.43) | 0.60 | 0.21 (0.03-1.77) | 0.15 |
| Owns chickens | 1.07 (0.34-3.37) | 0.91 | 3.86 (0.77-19.24) | 0.098 |
| Owns goats | 0.05 (0.01-0.41) | 0.005 | | |
| Owns pigs | 1.17 (0.37-3.69) | 0.79 | 1.78 (0.89-3.55) | 0.099 |
| Glass window covering | 1.37 (0.48-3.85) | 0.56 | 1.19 (0.34-4.23) | 0.78 |
| Travel in CMR in last 28 days | 2.6 (0.87-7.75) | 0.086 | 2.53 (0.78-8.16) | 0.12 |

**Supplemental Table 8.** ***Anopheles*** **abundance in Dschang neighborhoods**

| Health Area (number of households) | | Median (IQR) | Mean (SD) |
|---|---|---|---|
| **Bafou (n=4)** | *An. gambiae* | - | - |
| | *An. funestus* | - | - |
| | *An. nili* | - | - |
| | *An. ziemanni* | 0.5 (0-1.5) | 1 (1.41) |
| **Fialah-Foréké (n=25)** | *An. gambiae* | 0 (0-1) | 0.6 (1.26) |
| | *An. funestus* | - | - |
| | *An. nili* | - | - |
| | *An. ziemanni* | - | - |
| **Foto (n=35)** | *An. gambiae* | 1 (0-2) | 1.46 (2.59) |
| | *An. funestus* | - | - |
| | *An. nili* | - | - |
| | *An. ziemanni* | 0 (0-0) | 0.086 (0.37) |
| **Siteu (n=37)** | *An. gambiae* | 1 (0-3) | 3.24 (5.54) |
| | *An. funestus* | - | - |
| | *An. nili* | - | - |
| | *An. ziemanni* | - | - |

**Supplemental Figure 1. Distribution of parasitemias for *P. ovale spp*, *P. falciparum* and *P. vivax*.**

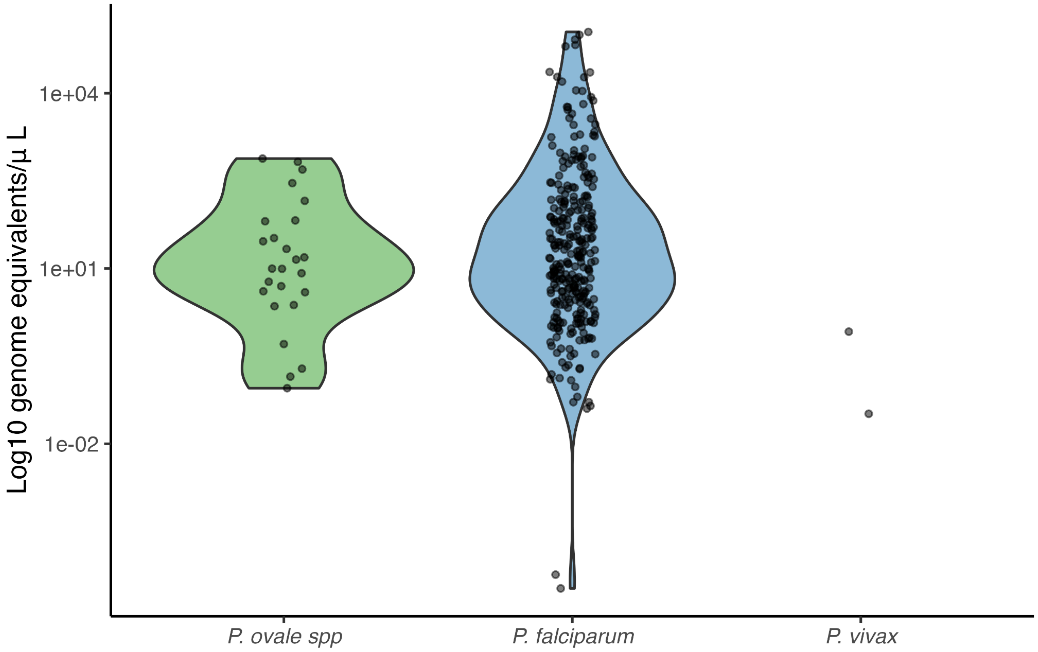

**Supplemental Figure 2. Correlation between gametocytemia and parasitemia in *P. falciparum*.**

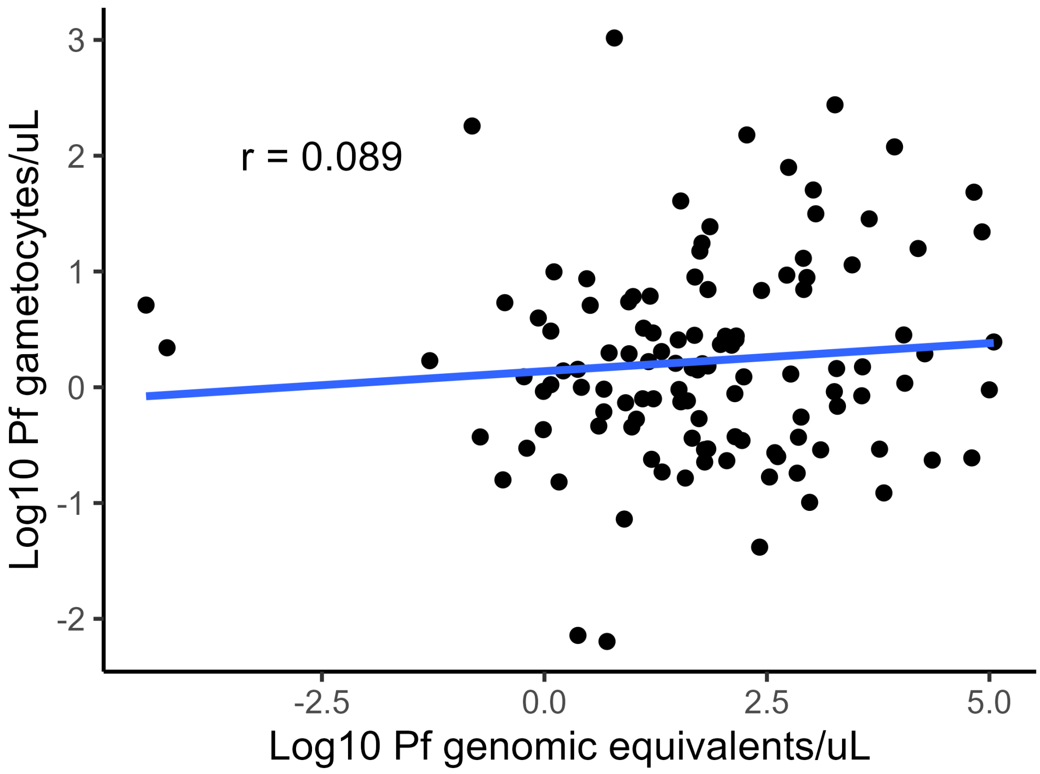

**Supplemental Figure 3. Limit of Detection (LOD) for *pomget* assay.**

A Probit analysis was conducted of replicate runs carried out using synthesized plasmids containing the *pomget* target sequence. An estimated LoD was calculated to be 281 transcript copies/uL (95% CI 162, 1310) based on the observed hit rate.

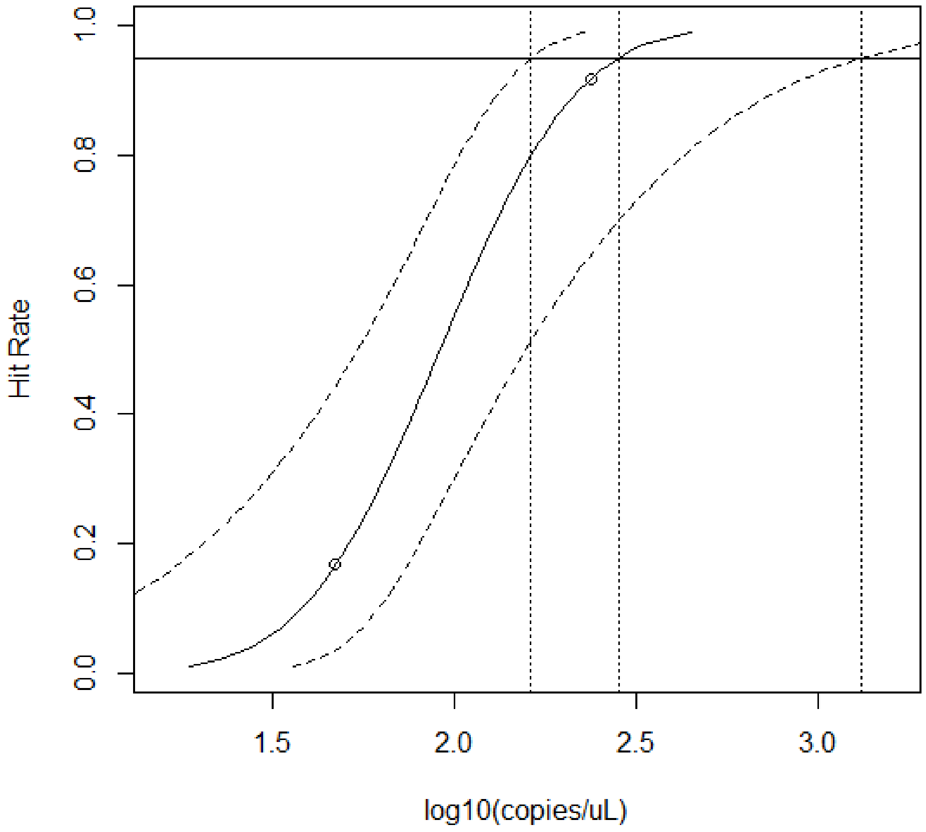

**Supplemental Figure 4. Mosquito capture time for indoor and outdoor collections across all households**

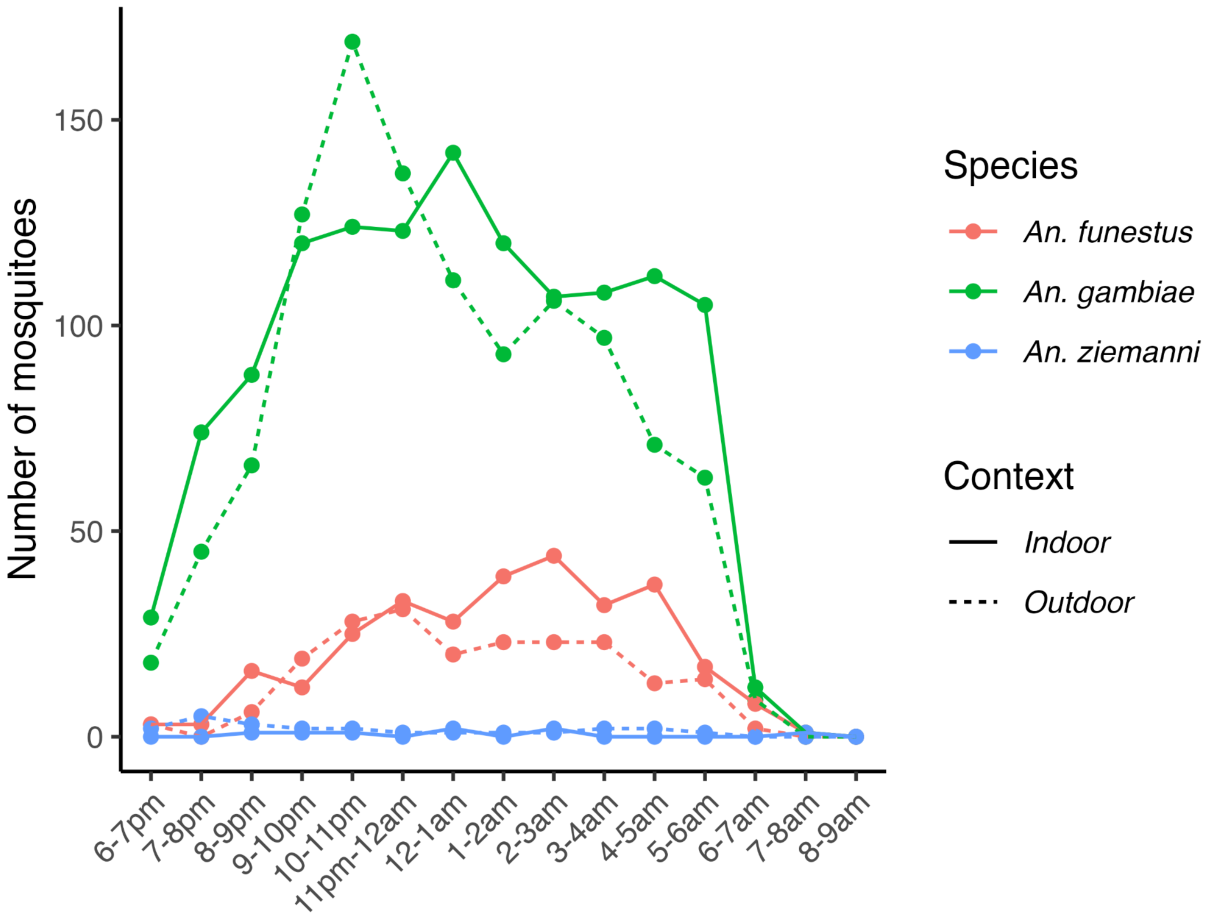
